# Multidomain Hypertension-Mediated Organ Damage in Ghanaian Adults: Prevalence, Predictors, and Implications for Cardiovascular Risk Stratification — A Secondary Analysis of the Ghana Heart Study

**DOI:** 10.64898/2026.05.21.26353747

**Authors:** K. O. Agyapong, E. Kyeremah, A. A. Folson, F. Agyekum, K. R. M. Blenman, L. T. Appiah, Y. Adu Boakye, I. K. Owusu

**Affiliations:** Yale School of Public Health, New Haven, CT, USA; Department of Medicine, Greater Accra Regional Hospital, Ridge-Accra, Ghana; Department of Epidemiology, Columbia University Mailman, School of Public Health, NY, USA; Department of Internal Medicine and Therapeutics, School of Medicine, University of Health and Allied Sciences, Ho, Ghana; Department of Internal Medicine, Ho Teaching Hospital, Ho, Ghana; Department of Medicine, College of Health Sciences, University of Ghana Medical School, University of Ghana, Accra, Ghana; Department of Medicine, School of Medicine and Dentistry, College of Health Sciences, Kwame Nkrumah University of Science and Technology, Kumasi, Ghana; Department of Internal Medicine, Section of Medical Oncology, Yale Cancer Center, New Haven, Connecticut, USA; Department of Computer Science, Yale School of Engineering and Applied Science, Yale University, New Haven, Connecticut, USA

**Author notes:** **Corresponding Author**: Kwabena Oteng Agyapong, MD, MPH, Yale School of Public Health, 60 College Street, New Haven, CT 06510, USA.

**Keywords:** hypertension-mediated organ damage, arterial stiffness, baPWV, target organ damage, Ghana, sub-Saharan Africa, cardiovascular risk stratification, pulse wave velocity, hypertension, left ventricular hypertrophy

## Abstract

**Background:** Comprehensive assessment of hypertension-mediated organ damage (HMOD) across multiple organ systems remains limited in sub-Saharan Africa. We aimed to determine the prevalence and predictors of multidomain HMOD in a geographically diverse Ghanaian adult population.

**Methods:** This secondary analysis of the Ghana Heart Study included 1,106 adults from four Ghanaian regions. Multidomain HMOD was defined as a pre-specified 9-domain composite score ≥2, constructed according to the ESH/ESC 2018 hypertension guidelines framework.^27^ Logistic regression and ROC analysis identified predictors and compared discriminative performance across vascular, cardiac, ECG, valvular, and renal domains.

**Results:** Mean age was 46.9 ± 17.2 years (58% female). Multidomain HMOD prevalence was 21.2% (235/1,106) and increased steeply with age: 8.6% in adults <45 years, 20.6% in those 45–59 years, and 44.4% in those ≥60 years. The HMOD-positive group was older (59.1 vs. 43.6 years), had higher systolic BP (147 vs. 123 mmHg), and higher hypertension prevalence (73% vs. 28%, all p<0.001). In adjusted models, the strongest associations were peripheral artery disease (OR 41.2, 95% CI 20.7–81.6), valvular burden (OR 14.4, 95% CI 4.8–43.8), and ECG-LVH (OR 9.0, 95% CI 5.9–13.8). baPWV demonstrated superior discriminative performance (AUC 0.827, 95% CI 0.794–0.860) compared with the ASCVD Pooled Cohort Equations (AUC 0.466; ΔAUC +0.351, DeLong test p<0.001). The metabolic syndrome triad was present in 13.4% and independently associated with multidomain HMOD (adjusted OR 2.11, 95% CI 1.28–3.49, p=0.004).

**Conclusions:** One in five Ghanaian adults has hypertension-mediated organ damage in ≥2 organ systems. baPWV is the strongest predictor and substantially improves risk stratification beyond conventional risk scores, supporting its use to guide hypertension management and HMOD assessment in West Africa.

## Introduction

Cardiovascular disease is the leading cause of death globally, with over 80% of mortality occurring in low- and middle-income countries.^1,2^ Sub-Saharan Africa bears a disproportionately high burden, with the age-standardized CVD prevalence in western SSA estimated at 9,475 per 100,000, the highest worldwide.^3^ Ghana, like many countries in the region, is undergoing a rapid epidemiological transition characterized by urbanization, dietary westernization, and increasing physical inactivity, all of which are driving a surge in hypertension and related complications.^4,5^ The Ghana Heart Study (2016–2017), a community-based cross-sectional survey of 1,106 adults from four regions, documented age-standardized prevalences of 15.1% for obesity, 26.1% for hypertension, 6.8% for diabetes mellitus, and 34.4% for dyslipidaemia.^6^ Behavioural risk factors were even more prevalent: 83.7% of participants were physically inactive, 81.4% had inadequate fruit intake, and 92% engaged in two or more unhealthy lifestyle behaviours.^7^

The concept of hypertension-mediated organ damage (HMOD), a subclinical structural and functional changes in the heart, blood vessels, kidneys, and other organs resulting from prolonged elevation of blood pressure, represents a critical intermediate stage in the cardiovascular disease continuum.^8^ Early detection of HMOD allows for intensified preventive efforts before irreversible end-organ failure develops. Previous analyses from the Ghana Heart Study reported single-organ HMOD prevalences of 10.1% for peripheral artery disease (PAD), 8.3% for carotid thickening, 4.1% for left ventricular hypertrophy (LVH), and 2.5% for chronic kidney disease (CKD).^6^ However, hypertension frequently causes damage across multiple organ systems simultaneously, and the burden of multidomain HMOD (involvement of two or more organ systems) has not been systematically evaluated in sub-Saharan African populations.

A major challenge for hypertension management in Ghana and broader West Africa is the poor performance of imported atherosclerotic cardiovascular disease (ASCVD) risk calculators. Agyekum et al.^9^ recently demonstrated only poor-to-moderate agreement among four commonly used tools (Pooled Cohort Equations, Framingham Risk Score, WHO/ISH charts, and Globorisk) in this population (kappa 0.3–0.8). The WHO/ISH tool, recommended by Ghana’s national CVD guidelines,^10^ classified 82% of adults aged 40–74 years as low risk, whereas the Framingham Risk Score classified 22% as high risk, a discrepancy with direct implications for decisions about blood pressure treatment intensity and intensification thresholds.

To our knowledge, this is among the first studies to apply the ESH/ESC 2018 framework for comprehensive multiorgan hypertension-mediated organ damage (HMOD) assessment in a sub-Saharan African community cohort and to evaluate the utility of brachial-ankle pulse wave velocity (baPWV) relative to conventional ASCVD risk scores in this setting.^27,28^

In this secondary analysis of the Ghana Heart Study, we aimed to: (1) determine the prevalence of multidomain HMOD in Ghanaian adults; (2) identify the strongest independent predictors of multidomain HMOD; (3) compare the discriminative performance of individual markers, particularly baPWV, against the ASCVD Pooled Cohort Equations for detecting multidomain HMOD; and (4) evaluate the incremental value of arterial stiffness measurement in hypertension risk stratification. These findings are interpreted together with previously published data on lifestyle risk factors^7^ and risk calculator performance^9^ from the same cohort, with the goal of informing targeted prevention strategies and the development of locally derived hypertension management models.

## Methods

### Study Design and Population

This is a secondary analysis of the Ghana Heart Study, a community-based cross-sectional survey conducted between September 2016 and March 2017. The parent study used a 3-stage stratified random sampling strategy to recruit 1,106 adults aged ≥18 years from eight communities (four urban, four rural) across four geographically diverse regions of Ghana: Greater Accra, Ashanti, Central, and Northern. Detailed methodology of the parent study has been published previously.^6,7^

Exclusion criteria included pregnancy, type 1 diabetes mellitus, established atherosclerotic cardiovascular disease (self-reported stroke, myocardial infarction, or peripheral artery disease), congenital heart disease, secondary hypertension, or refusal to provide informed consent. The study was registered at ChiCTR1800017374 and received ethical approval from the Committee on Human Research, Publications and Ethics of the Kwame Nkrumah University of Science and Technology and Komfo Anokye Teaching Hospital (CHRPE/AP/415/16). Written informed consent was obtained from all participants.

### Data Collection

All data were collected during the original Ghana Heart Study using standardized protocols by trained research assistants (medical officers and nurses), as previously described in detail.^6,7^

### Demographic and lifestyle variables

Age, sex, ethnicity, education, marital status, employment status, region, and urban/rural residence were recorded. Smoking was defined as current use of any tobacco product (daily or occasionally). Physical inactivity was defined as <150 minutes of moderate-to-vigorous physical activity per week. Inadequate fruit and vegetable intake was defined as <2 servings per day, following WHO STEPwise approach criteria.^7,11^

### Anthropometric and blood pressure measurements

Weight, height, and waist circumference were measured with participants in light clothing and bare feet. Body mass index (BMI) was calculated as weight(kg)/height(m)². Blood pressure was measured three times using an OMRON M6 device after 10 minutes of rest, with the average of the last two readings used for analysis. Hypertension was defined as systolic BP ≥140 mmHg and/or diastolic BP ≥90 mmHg or current use of antihypertensive medication.^6^

### Laboratory measurements

Fasting venous blood samples (8–12 hours overnight fast) were analyzed for total cholesterol, LDL cholesterol, HDL cholesterol, triglycerides, HbA1c, uric acid, hs-CRP, and creatinine. Dyslipidaemia was defined as TG ≥1.70 mmol/L, TC ≥6.22 mmol/L, LDL ≥4.14 mmol/L, HDL <1.04 mmol/L, use of lipid-lowering medications, or lipoprotein(a) ≥60 mg/dL.^6^

### Vascular Measurements

#### Arterial stiffness (baPWV)

Brachial-ankle pulse wave velocity was measured using an OMRON/Colin VP-2000 automated oscillometric device.^21^ Measurements were taken from both limbs, and the mean of left and right values was used (or the single available value if only one side was valid). baPWV values are reported in meters per second (m/s). High baPWV was defined as ≥14 m/s based on the distribution in this cohort and prior literature.

Peripheral artery disease (PAD) was defined as ankle-brachial index (ABI) <0.9 in either leg, measured using the same VP-2000 device. Carotid intima-media thickness (IMT) was measured by ultrasound (GE VIVID Q, 9L-RS probe). All IMT values reported in this manuscript represent the true intima-media thickness (mean 0.80 mm overall), not the outer vessel diameter. IMT data were filtered to exclude sentinel values (-1) and outliers (>2.0 mm). Carotid plaque (present/absent) was also recorded.^6^

### Echocardiographic Measurements

Transthoracic echocardiography (GE VIVID Q, M4S-RS probe) was performed by trained cardiologists. Left ventricular mass index (LVMI) was calculated using the Devereux formula and indexed to body surface area. Echocardiographic LVH was defined as LVMI >95 g/m² in women and >115 g/m² in men.^6,12^ Left ventricular geometry was classified as normal, concentric remodelling (relative wall thickness [RWT] ≥0.42, normal LVMI), eccentric LVH (increased LVMI, RWT <0.42), or concentric LVH (increased LVMI, RWT ≥0.42).

#### Diastolic function assessment (ASE/EACVI 2016 criteria)

Diastolic dysfunction was diagnosed according to the 2016 ASE/EACVI recommendations ^13^, requiring ≥2 of the following three binary parameters:

1. Septal e’ velocity <7 cm/s OR lateral e’ velocity <10 cm/s (tissue Doppler imaging)
2. Average E/E’ ratio >14
3. Left atrial diameter >40 mm (or left atrial volume index >34 mL/m²)

E/A ratio was used only for grading the severity of dysfunction (Grade I = impaired relaxation, E/A <0.8; Grade II = pseudonormal, E/A 0.8–2.0; Grade III = restrictive, E/A >2.0) and was not included as a diagnostic criterion for the presence of diastolic dysfunction.

Valvular regurgitation was graded 0–4 (absent, trace, mild, moderate, severe), with significant regurgitation defined as grade ≥3.

### ECG Measurements

ECG-LVH was defined as Sokolow-Lyon (SV1 + RV5/V6 ≥3.5 mV) or Cornell (RaVL + SV3 >2.8 mV in men, >2.0 mV in women) criteria.

### Renal Measurements

Estimated glomerular filtration rate (eGFR) was calculated using the CKD-EPI equation. CKD stage ≥3 was defined as eGFR <60 mL/min/1.73m². Urine albumin-to-creatinine ratio (UACR) was measured, with microalbuminuria defined as 30–299 mg/g and macroalbuminuria as ≥300 mg/g.

### Definition of Multidomain Hypertension-Mediated Organ Damage (HMOD)

We constructed a 9-domain composite score of hypertension-mediated organ damage (HMOD) based on the ESH/ESC 2018 hypertension guidelines framework^27^ and established pathophysiological links to hypertension-mediated organ injury. Each domain was coded as present (1) or absent (0):

1. High baPWV (≥14 m/s) — vascular stiffness
2. Peripheral artery disease (ABI ≤0.9) — peripheral atherosclerosis
3. Carotid plaque — carotid atherosclerosis
4. ECG-LVH (Sokolow-Lyon or Cornell criteria) — electrical remodelling
5. Echocardiographic LVH (LVMI >115 g/m² men / >95 g/m² women) — structural cardiac damage
6. Diastolic dysfunction (ASE/EACVI 2016 criteria) — functional cardiac impairment
7. Significant valvular regurgitation (grade ≥3, moderate–severe) — valvular damage
8. CKD stage ≥3 (eGFR ≤60 mL/min/1.73m²) — renal impairment
9. Albuminuria (UACR ≥30 mg/g) — renal microvascular damage

Multidomain TOD was pre-specified as a composite score ≥2, reflecting involvement of at least two distinct organ systems. This threshold was chosen because multi-organ involvement is clinically meaningful and has been used in prior European studies of hypertension-mediated organ damage ^28^. A sensitivity analysis using a more stringent threshold (≥3 domains) was performed to assess robustness.

#### Justification of the Multidomain TOD Composite

Individual subclinical markers (e.g., LVH, arterial stiffness, microalbuminuria) are well-validated surrogates for future cardiovascular events in multiple populations.^8,27^ By requiring involvement of ≥2 domains, our definition captures the systemic nature of hypertension-mediated injury and identifies individuals with more advanced subclinical disease. This approach aligns with the ESH/ESC 2018 recommendation to assess damage across multiple organs for comprehensive risk stratification.^27^ While the composite has not yet been prospectively validated against hard cardiovascular events in sub-Saharan African populations, each component has strong biological plausibility and established prognostic value. Future longitudinal studies will be essential to confirm the composite’s predictive value for clinical outcomes.

### ASCVD Risk Score Calculation

For participants aged 40–74 years (n=615), we calculated 10-year ASCVD risk using the Pooled Cohort Equation (PCE) ^14^ as the reference standard, given its inclusion of Black populations in its derivation cohorts.

### Statistical Analysis

Analyses were performed using R version 4.3.3. Continuous variables were expressed as mean (SD) or median (IQR) and compared using Wilcoxon rank-sum test or Kruskal-Wallis test. Categorical variables were expressed as n (%) and compared using Pearson’s chi-square test or Fisher’s exact test.

#### Logistic regression

Multivariable logistic regression was used to identify independent predictors of multidomain TOD, adjusting for age, systolic BP, BMI, dyslipidaemia, and smoking. Results are reported as odds ratios (OR) with 95% confidence intervals (CI).

#### ROC analysis

Area under the receiver operating characteristic curve (AUC) was calculated for each candidate marker (baPWV, LVMI, eGFR, E/E’, ABI, IMT, hs-CRP, RVSP, EF, and the ASCVD risk score) to discriminate multidomain TOD status. AUC values were compared using DeLong’s test.

#### Incremental value

The incremental value of adding baPWV to the ASCVD risk score was assessed by comparing AUC (DeLong test) and model fit (likelihood ratio test) (see Table S3).

#### Missing data

Missingness was quantified for each variable. Given low missingness for most variables (baPWV 1.6%, ABI 1.7%, E/E’ 8.3%), complete-case analysis was performed. UACR had higher missingness (36.8%); analyses involving UACR are reported separately with denominators stated. Missing baPWV data (n=18, 1.6%) were confirmed as missing completely at random by Little’s MCAR test (p=0.14 for age, p=1.00 for sex; borderline association with hypertension [p=0.03] noted as a limitation). The primary logistic model included 11 covariates with 235 events (events per variable = 21.4), satisfying conventional EPV thresholds (recommended minimum ≥10). A p-value <0.05 (two-sided) was considered statistically significant.

## Results

### Participant Characteristics

A total of 1,106 participants were included. The mean age was 46.9 (17.2) years, and 58.0% were female. The majority (80.4%) resided in urban areas, and 57.8% were of Akan ethnicity (Table 1).

**Table 1.**
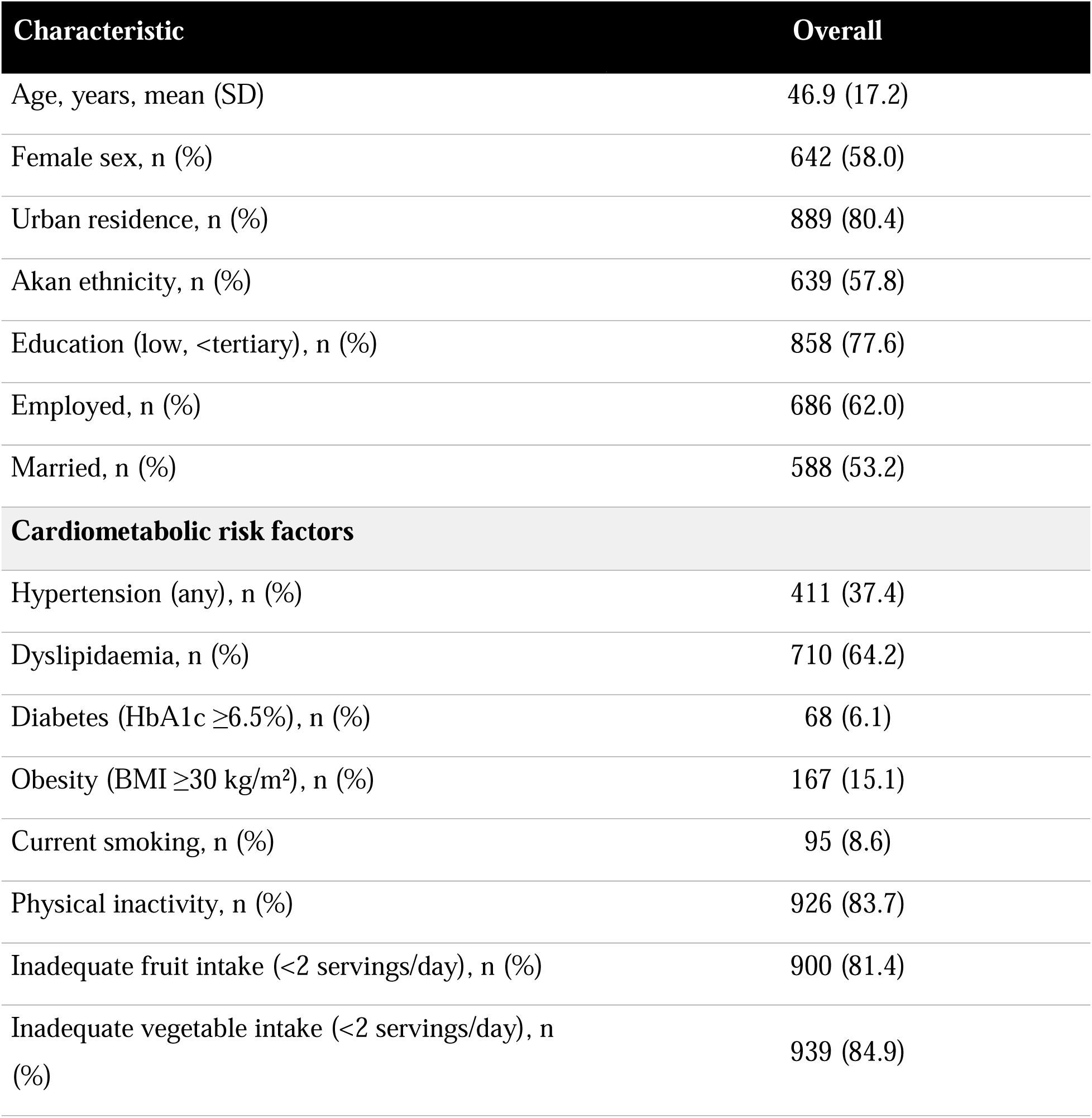
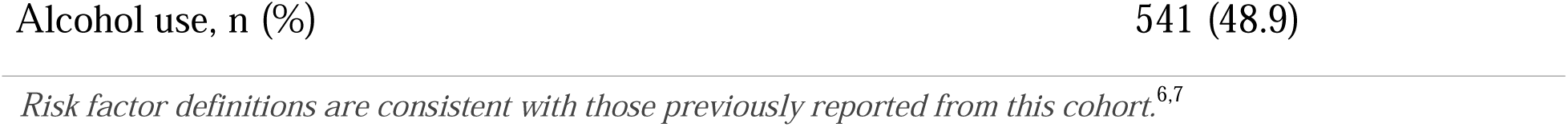
Baseline Characteristics of the Study Population (N=1,106)

### Prevalence of Multidomain Target Organ Damage

The composite TOD score distribution is shown in Figure 1 and Table S1. Multidomain TOD (composite score ≥2) was present in 235 participants (21.2%). Among those with multidomain TOD, the majority had a score of 2 (66.0%) or 3 (24.7%), with only 2.0% having scores of 4 or 5.

**Figure 1.**
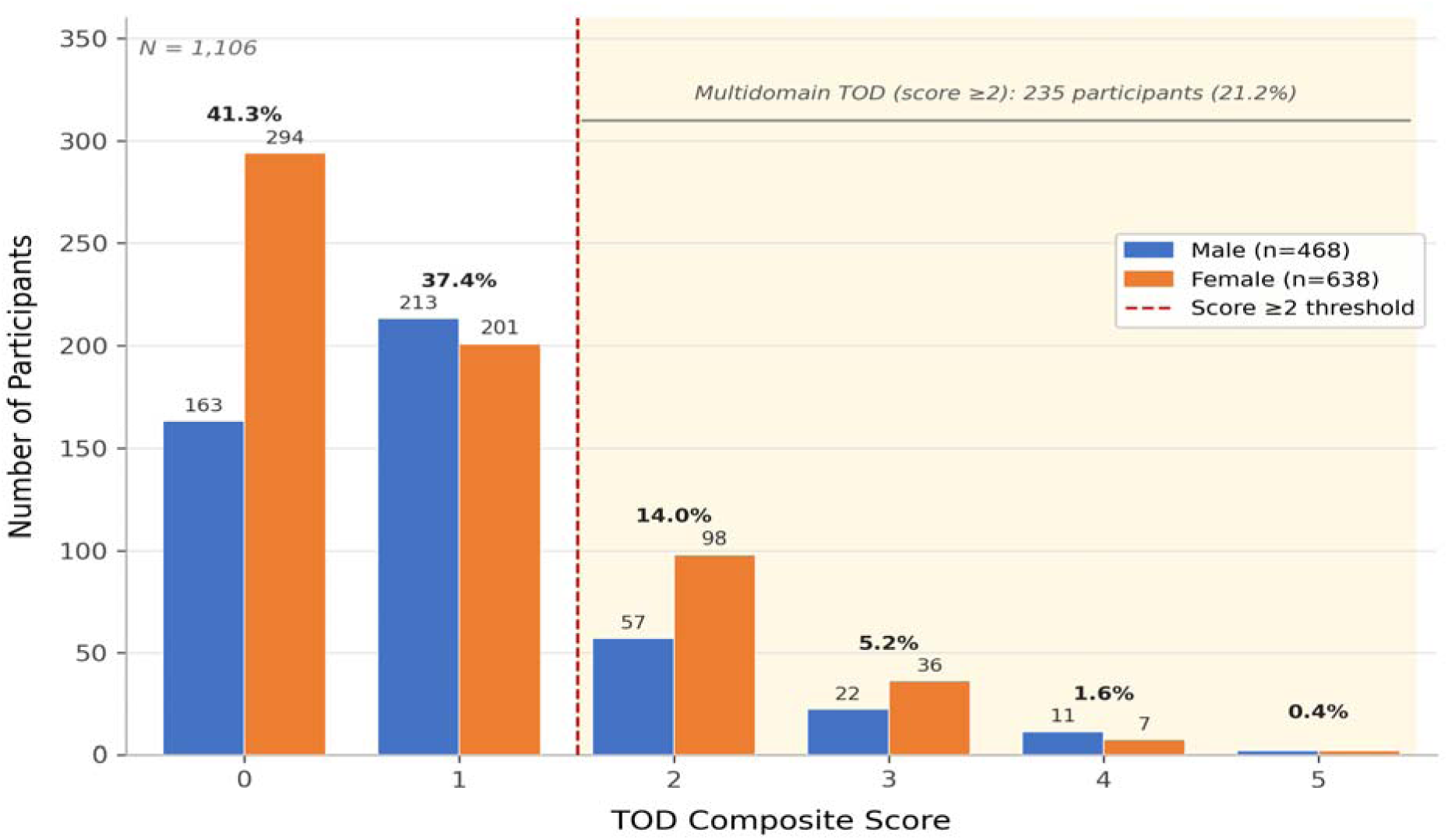
Distribution of the TOD Composite Score. Distribution of the 9-domain TOD composite score in 1,106 Ghanaian adults. Most participants had a score of 0 (41.3%) or 1 (37.4%), while 21.2% had multidomain TOD (composite score ≥2). The distribution demonstrates that a substantial proportion of the population has evidence of subclinical damage across multiple organ systems. *TOD = target organ damage (composite score). Multidomain HMOD defined as composite score ≥2 (TOD composite score)*.

### Characteristics by Multidomain TOD Status

Participants with multidomain TOD were significantly older (59.1 (16.8) vs. 43.6 (15.8) years, p < 0.001) and had higher systolic blood pressure (147.1 (26.7) vs. 123.2 (18.9) mmHg, p < 0.001). Hypertension prevalence was nearly threefold higher in the TOD group (73% vs. 28%, p < 0.001). Dyslipidaemia was also more common (71% vs. 62%, p = 0.02). Complete characteristics across all domains are presented in Table 2.

**Table 2.** Participant Characteristics by Multidomain TOD Status.

| Domain / Parameter | No TOD (n=871) | TOD (n=235) | p-value |
| --- | --- | --- | --- |
| **Demographics** |  |  |  |
| Age, years, mean (SD) | 43.6 (15.8) | 59.1 (16.8) | <0.001 |
| Male sex, n (%) | 374 (43) | 90 (38) | 0.2 |
| **Anthropometrics** |  |  |  |
| BMI, kg/m<sup>2</sup>, mean (SD) | 25.7 (5.8) | 26.4 (6.1) | 0.064 |
| Central obesity (waist >102/88 cm), n (%) | 278 (35) | 102 (44) | 0.006 |
| **Blood Pressure** |  |  |  |
| Systolic BP, mmHg, mean (SD) | 123.2 (18.9) | 147.1 (26.7) | <0.001 |
| Diastolic BP, mmHg, mean (SD) | 76.7 (11.5) | 86.9 (16.1) | <0.001 |
| Hypertension (any definition), n (%) | 240 (28) | 171 (73) | <0.001 |
| **Metabolic Risk Factors** |  |  |  |
| Dyslipidaemia, n (%) | 544 (62) | 166 (71) | 0.02 |
| Diabetes (HbA1c $\geq 6.5\%$), n (%) | 41 (4.8) | 27 (10.5) | 0.009 |
| **Vascular** |  |  |  |
| baPWV, m/s, mean (SD) | 11.4 (2.2) | 14.9 (3.0) | <0.001 |
| High baPWV ($\geq 14$ m/s), n (%) | 96 (11.0) | 139 (59.1) | <0.001 |
| ABI (minimum), mean (SD) | 1.02 (0.07) | 0.96 (0.09) | <0.001 |
| PAD (ABI<0.9), n (%) | 29 (3.3) | 66 (28.1) | <0.001 |
| Carotid IMT (true CIMT), mm, mean (SD) | 0.76 (0.23) | 0.95 (0.21) | <0.001 |
| Carotid plaque, n (%) | 2 (0.2) | 20 (8.5) | <0.001 |
| **ECG** |  |  |  |
| ECG-LVH (Sokolow-Lyon $\geq 1$ mV), n (%) | 270 (31) | 166 (71) | <0.001 |
| ECG-LVH (Cornell $\geq 1$ mV), n (%) | 44 (5.1) | 55 (23) | <0.001 |
| ECG-LVH (either criterion), n (%) | 284 (33) | 186 (79) | <0.001 |
| **Echocardiographic — Structure** |  |  |  |
| LVMI, g/m<sup>2</sup>, mean (SD) | 69.0 (15.2) | 83.9 (26.3) | <0.001 |
| Echo-LVH, n (%) | 7 (0.9) | 58 (25.0) | <0.001 |
| LV geometry, n (%) |  |  | <0.001 |
| — Normal | 524 (66) | 69 (30) |  |
| — Concentric remodelling | 261 (33) | 103 (45) |  |
| — Eccentric LVH | 1 (0.1) | 15 (6.5) |  |
| — Concentric LVH | 6 (0.8) | 43 (19) |  |
| LA enlargement (LAD >40 mm), n (%) | 119 (15) | 122 (53) | <0.001 |
| **Echocardiographic — Systolic Function** |  |  |  |
| Ejection fraction, %, mean (SD) | 65.9 (5.1) | 65.6 (6.0) | 0.8 |
| S' average TDI, cm/s, mean (SD) | 8.4 (1.6) | 7.8 (1.6) | <0.001 |
| Low S' (<7.5 cm/s), n (%) | 224 (26) | 100 (43) | <0.001 |
| **Echocardiographic — Diastolic Function** |  |  |  |
| E/A ratio, mean (SD) | 1.5 (5.0) | 0.9 (0.5) | <0.001 |
| Impaired relaxation (E/A <0.8), n (%) | 135 (17) | 133 (58) | <0.001 |
| E/E' average, mean (SD) | 7.4 (2.0) | 9.2 (3.4) | <0.001 |
| Elevated E/E' (>14), n (%) | 2 (0.2) | 19 (8.1) | <0.001 |
| Low E' (septal <7 or lateral <10), n (%) | 229 (26) | 173 (74) | <0.001 |
| Diastolic dysfunction (ASE 2016 criteria), n (%) | 2 (0.2) | 27 (11) | <0.001 |
| **Right Heart** |  |  |  |
| RVSP, mmHg, mean (SD) | 11.2 (0.5) | 11.1 (0.4) | <0.001 |
| Pulmonary hypertension (RVSP >36), n (%) | 1 (0.1) | 3 (1.3) | 0.03 |
| **Valvular** |  |  |  |
| Any significant regurgitation (≥moderate), n (%) | 6 (0.7) | 16 (6.8) | <0.001 |
| Valvular burden score, n (%) |  |  | <0.001 |
| — 0 | 865 (99) | 219 (93) |  |
| — 1 | 6 (0.7) | 13 (5.5) |  |
| — 2 | 0 (0) | 3 (1.3) |  |
| **Renal** |  |  |  |
| eGFR, mL/min/1.73m<sup>2</sup>, mean (SD) | 85.4 (7.0) | 77.5 (15.5) | <0.001 |
| CKD stage $\geq 3$ (eGFR<60), n (%)* | 2 (0.3) | 34 (14.5) | <0.001 |
| UACR, mg/g, mean (SD) | 1.9 (4.9) | 12.2 (51.3) | <0.001 |
| Microalbuminuria (30-299 mg/g), n (%)* | 3 (0.4) | 12 (5.1) | <0.001 |
| **Composite Scores** |  |  |  |
| ASCVD 10-year risk, %, mean (SD) | 22.6 (27.7) | 23.3 (24.9) | 0.13 |
| High ASCVD risk ($\geq 7.5\%$), n (%) | 387 (44) | 117 (50) | 0.2 |
| Composite Z-score, mean (SD) | -0.2 (0.5) | 0.6 (0.7) | <0.001 |

Data are mean (SD) or n (%). TOD = target organ damage (score $\geq 2$ ). eGFR measured in 915 participants; CKD stage $\geq 3$ present in 36 participants (3.3% of total cohort; 3.9% of those with eGFR measured). UACR measured in 699 participants. IMT measured in 1,058 participants (870 No TOD, 188 TOD). Valvular burden score = number of valves with grade $\geq 3$ (moderate–severe) regurgitation (range 0–2); significant regurgitation was defined as any valve graded $\geq 3$ , consistent with ESH/ESC 2018 guidelines<sup>27</sup>.

### Adjusted Odds Ratios for Multidomain Target Organ Damage

Multivariable logistic regression adjusting for age, systolic BP, BMI, dyslipidaemia, and smoking revealed the strongest independent associations with multidomain TOD (Figure 2; Table 3). PAD (ABI<0.9) had the highest OR (41.2, 95% CI 20.7–81.6), followed by valvular burden (OR 14.4, 95% CI 4.8–43.8) and ECG-LVH (OR 9.0, 95% CI 5.9–13.8). baPWV was also significantly associated (OR 1.44 per m/s, 95% CI 1.29–1.60, p<0.001). In contrast, traditional metabolic markers (LDL, HbA1c, hs-CRP) were not significant after adjustment.

**Figure 2.**
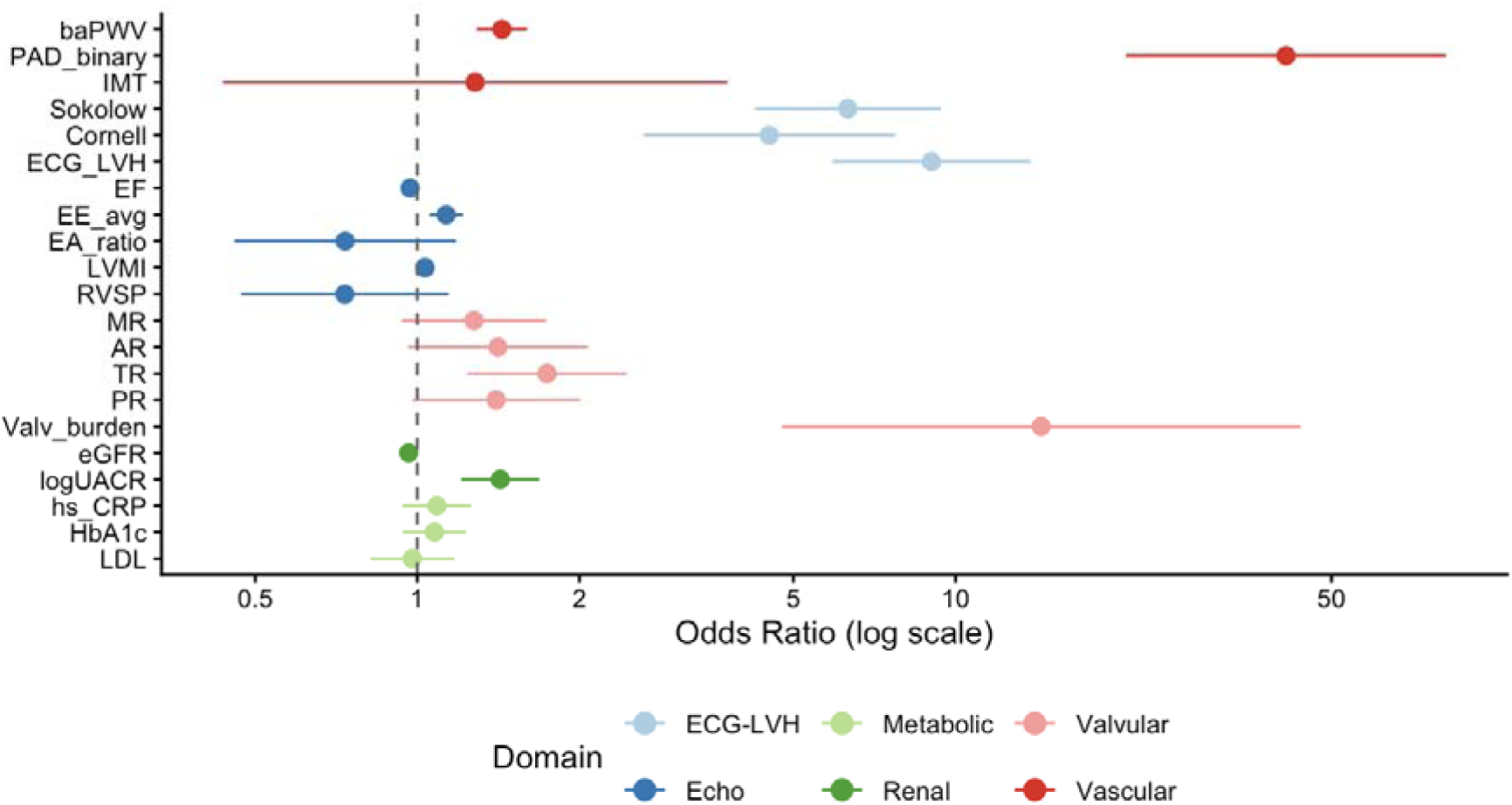
Adjusted Odds Ratios for Multidomain Hypertension-Mediated Organ Damage. Forest plot showing multivariable-adjusted odds ratios (95% CI) for independent predictors of multidomain HMOD. Models were adjusted for age, systolic blood pressure, BMI, dyslipidaemia, and smoking. ORs are displayed on a log scale and colour-coded by organ domain. PAD, valvular burden score, and ECG-LVH were the strongest predictors. *PAD = peripheral artery disease; ABI = ankle-brachial index; ECG-LVH = electrocardiographic left ventricular hypertrophy; OR = odds ratio; CI = confidence interval; TOD = target organ damage*.

**Table 3.** Adjusted Odds Ratios for Multidomain Target Organ Damage.

| **Predictor** | **OR (95% CI)** | **p-value** |
| --- | --- | --- |
| **Vascular** |  |  |
| PAD (ABI<0.9) | 41.15 (20.75 – 81.64) | <0.001 |
| baPWV (per m/s) | 1.44 (1.29 – 1.60) | <0.001 |
| **ECG-LVH** |  |  |
| ECG-LVH (any criterion) | 9.03 (5.90 – 13.81) | <0.001 |
| **Valvular** |  |  |
| Valvular burden (per grade) | 14.43 (4.76 – 43.79) | <0.001 |
| **Renal** |  |  |
| log(UACR) (per unit) | 1.43 (1.21 – 1.69) | <0.001 |
| eGFR (per mL/min/1.73m<sup>2</sup>) | 0.96 (0.95 – 0.98) | <0.001 |
| **Echocardiographic** |  |  |
| E/E' (per unit) | 1.13 (1.05 – 1.22) | 0.001 |
| LVMI (per g/m<sup>2</sup>) | 1.03 (1.02 – 1.04) | <0.001 |
| **Metabolic (not significant)** |  |  |
| LDL cholesterol | 0.98 (0.82 – 1.17) | 0.82 |
| HbA1c | 1.08 (0.94 – 1.23) | 0.29 |
| hs-CRP (log) | 1.09 (0.94 – 1.26) | 0.27 |

*Adjusted for age, systolic BP, BMI, dyslipidaemia, and smoking.*

### Discriminative Performance of Individual Markers

ROC analysis showed that baPWV had the best discriminative performance among non-composite markers for detecting multidomain HMOD (AUC 0.827, 95% CI 0.794–0.860) (Figure 3). baPWV demonstrated superior discriminative compared with the ASCVD Pooled Cohort Equations (AUC 0.466; ΔAUC +0.351, DeLong test p<0.001). baPWV outperformed LVMI (0.670), eGFR (0.646), E/E’ (0.670), and the ASCVD risk score (0.466) (Table 4). The incremental value of adding baPWV to the ASCVD risk score is detailed in Table S3.

**Figure 3.**
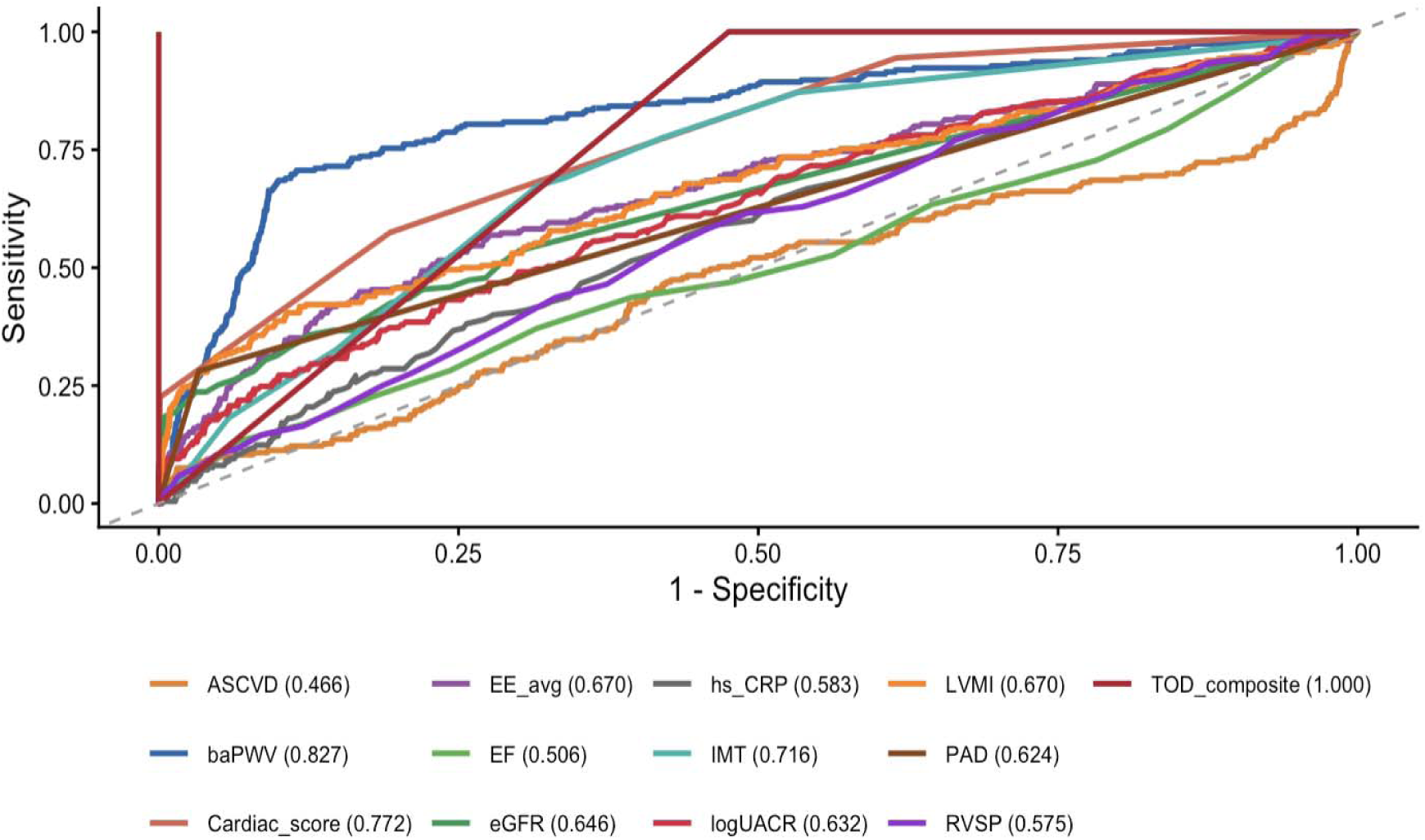
Receiver Operating Characteristics (ROC) Curves for Prediction of Multidomain Target Organ Damage — All Markers vs Multidomain TOD. ROC curves comparing the discriminative performance of baPWV, cardiac TOD score, carotid IMT, E/E’ ratio, LVMI, eGFR, and the ASCVD Pooled Cohort Equations for detecting multidomain TOD (composite score ≥2). baPWV achieved the highest AUC (0.827, 95% CI 0.794–0.860), substantially outperforming the ASCVD risk score (AUC 0.466; ΔAUC +0.351, DeLong test p<0.001). The TOD composite score is shown for reference only (tautological, AUC 1.000). *AUC = area under the curve; baPWV = brachial-ankle pulse wave velocity; IMT = intima-media thickness; LVMI = left ventricular mass index; ASCVD = atherosclerotic cardiovascular disease; TOD = target organ damage*.

**Table 4.** Area Under the Curve (AUC) for Multidomain TOD.

| Marker | AUC | 95% CI |
| --- | --- | --- |
| TOD Composite Score | 1.000 | 1.000 – 1.000 |
| **baPWV** | **0.827** | 0.794 – 0.860 |
| Cardiac TOD Score | 0.772 | 0.741 – 0.803 |
| Carotid IMT (true CIMT) | 0.716 | 0.676 – 0.755 |
| E/E' average | 0.670 | 0.627 – 0.713 |
| LVMI | 0.670 | 0.626 – 0.714 |
| eGFR | 0.646 | 0.605 – 0.688 |
| log(UACR) | 0.632 | 0.582 – 0.681 |
| PAD (binary) | 0.624 | 0.594 – 0.653 |
| hs-CRP (log) | 0.583 | 0.539 – 0.627 |
| RVSP | 0.575 | 0.532 – 0.618 |
| Ejection fraction | 0.506 | 0.460 – 0.553 |
| ASCVD 10-year risk (PCE) | 0.466 | 0.418 – 0.515 |

### Sensitivity Analysis

A sensitivity analysis using a stricter definition of multidomain TOD (composite score ≥3) yielded consistent findings. Prevalence was 7.2% (80/1,106), and baPWV remained a strong predictor (OR per SD 1.77, 95% CI 1.20–2.62, p=0.004; AUC 0.874). Full results are presented in Table S2.

### Age Group Analysis

The prevalence of multidomain TOD increased sharply and progressively with age (Table 5 and Figure 4). Prevalence was 8.6% in adults <45 years, 20.6% in those 45–59 years, and 44.4% in those ≥60 years. The number needed to screen (NNS) to detect one case of multidomain TOD was 12, 5, and 2 in these respective age groups.

**Figure 4.**
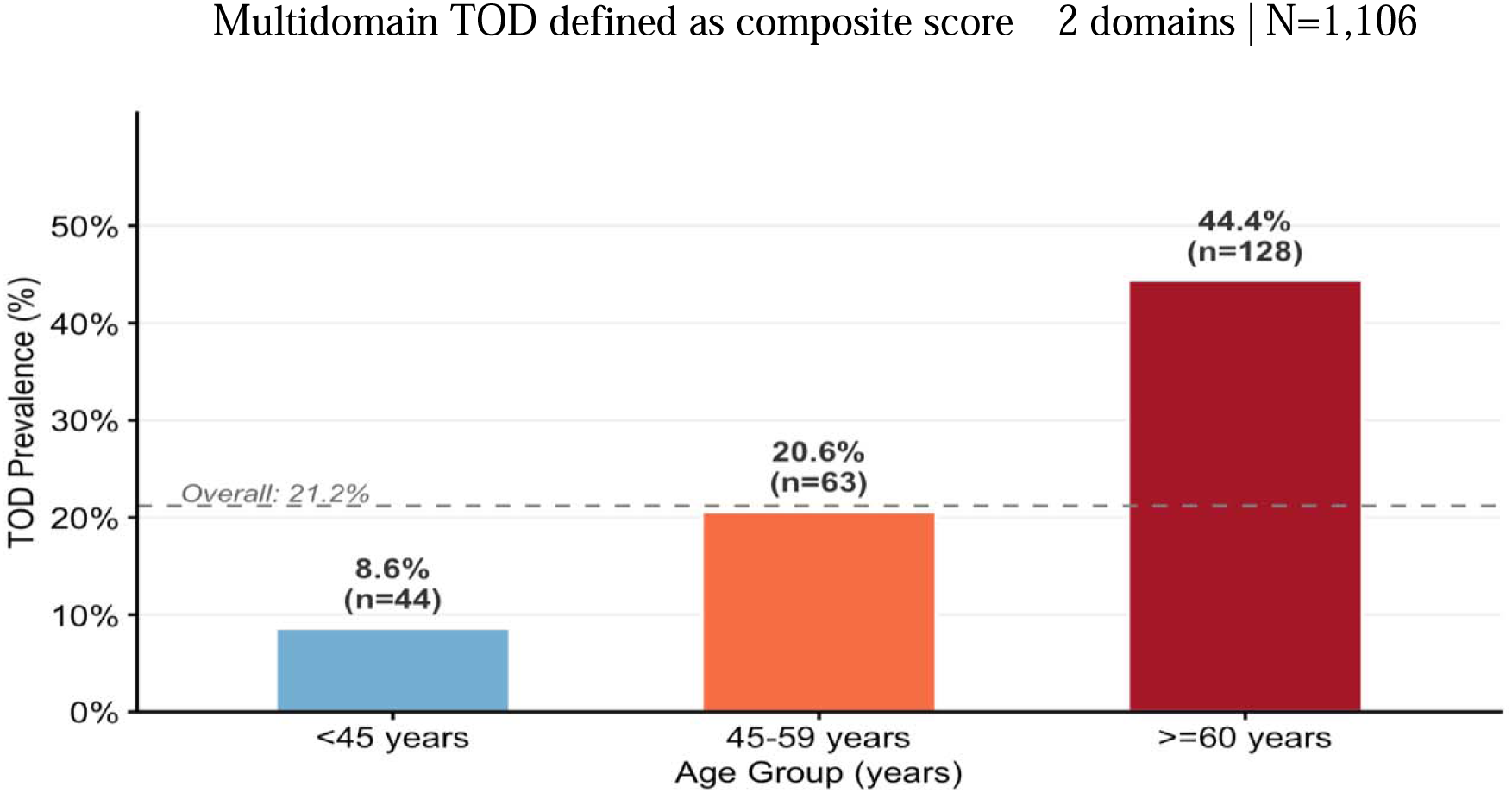
Prevalence of Multidomain Hypertension-Mediated Organ Damage by Age Group. Multidomain TOD defined as composite score 2 domains | N=1,106 Age-stratified prevalence of multidomain HMOD among 1,106 Ghanaian adults. Prevalence increased markedly with age: 8.6% (≤45 years), 20.6% (45–59 years), and 44.4% (≥60 years). Numbers inside bars indicate the number needed to screen (NNS) to detect one case. Dashed horizontal line represents the overall prevalence (21.2%). *TOD = target organ damage; NNS = number needed to screen*.

**Table 5.** Prevalence of Multidomain Hypertension-Mediated Organ Damage by Age Group.

| Age Group | Total N | Multidomain<br>TOD n | Prevalence<br>(%) | 95% CI* | NNS† |
| --- | --- | --- | --- | --- | --- |
| <45 years | 512 | 44 | 8.6% | 6.3 – 11.4 | 12 |
| 45-59 years | 306 | 63 | 20.6% | 16.3 – 25.5 | 5 |
| ≥60 years | 288 | 128 | 44.4% | 38.7 – 50.3 | 2 |
| **All ages** | **1,106** | **235** | **21.2%** | **18.9 – 23.7** | **5** |

\*95% confidence intervals calculated using the Wilson score method. †Number Needed to Screen = 1 / prevalence (rounded to nearest whole number).

### Additional Findings

Cardiometabolic triad analyses are presented in Table S4. The metabolic syndrome triad (central obesity + hypertension + dyslipidaemia or diabetes) was present in 148 participants (13.4%; adjusted OR 2.11, 95% CI 1.28–3.49, p = 0.004). Rarer triads (vascular-renal and cardio-renal) were uncommon (fewer than 0.5%).

## Discussion

This study provides one of the first comprehensive assessments of hypertension-mediated organ damage (HMOD) across multiple organ systems in a West African community-based cohort and applies the ESH/ESC 2018 multiorgan HMOD framework in sub-Saharan Africa.^27^

The principal finding is that baPWV is the strongest single predictor of multidomain HMOD (AUC 0.827), markedly outperforming the ASCVD Pooled Cohort Equations (AUC 0.466; ΔAUC +0.351, DeLong test p<0.001). In a population where hypertension drives 73% of HMOD cases, this superior performance demonstrates that baPWV can identify hypertension-mediated injury that conventional risk scores miss entirely and supports its use to guide intensification of antihypertensive therapy in resource-limited settings.

The strongest independent associations were observed with peripheral artery disease (OR 41.2), valvular burden score (OR 14.4), and ECG-LVH (OR 9.0). Prevalence of multidomain HMOD rose steeply with age, from 8.6% in adults <45 years to 44.4% in those ≥60 years, yielding a number needed to screen of only 2 in the oldest group, underscoring the cumulative effect of long-standing hypertension and the efficiency of age-targeted screening.

A striking finding was the sevenfold discrepancy between ECG-LVH (42% overall, 79% in the HMOD group) and echocardiographic LVH (6.4% overall, 25% in the HMOD group). This pattern, well described in Black African populations,^18,19^ has important implications for hypertension staging and clinical practice, as ECG voltage criteria have limited specificity for true structural LVH in this demographic and may lead to over-diagnosis if used in isolation.

The metabolic syndrome triad was present in 13.4% of participants and remained independently associated with multidomain HMOD (adjusted OR 2.11, 95% CI 1.28–3.49). In contrast, vascular-renal and cardio-renal triads were rare (fewer than 0.5%), consistent with the low overall prevalence of CKD stage ≥3 (3.3%), although CKD was present in 14.5% of the HMOD group, reflecting the younger age structure of this cohort.

### Comparison with Previous Findings from the Ghana Heart Study

Our findings extend and complement previously published work from the same cohort. Li et al.^6^ reported single-organ HMOD prevalences of 10.1% for PAD, 8.3% for carotid thickening, 4.1% for LVH, and 2.5% for CKD. Our finding that 21.2% have multidomain HMOD indicates that single-organ estimates substantially underestimate the true burden of hypertension-mediated injury. Our multivariable model confirms hypertension as central but additionally identifies PAD (OR 41.2), valvular burden (OR 14.4), and ECG-LVH (OR 9.0) as the strongest independent predictors of multidomain HMOD.

Agyekum et al.^7^ documented that 92% of participants engage in two or more unhealthy lifestyle behaviours, with physical inactivity (84%) and inadequate fruit/vegetable intake (81–85%) being most common. The high prevalence of multidomain HMOD (21%) is consistent with this lifestyle risk factor burden. Higher education was protective (OR 0.3, p=0.01),^7^ whereas employment was associated with increased risk (OR 5.4, p=0.003), socio-demographic gradients that should inform targeted prevention strategies.

### Diastolic Function Assessment in Hypertensive Adults

Using the ASE/EACVI 2016 criteria,^13^ definite diastolic dysfunction was present in only 2.6% of the overall cohort but 11% of the HMOD group. A much larger proportion (24% overall, 58% in the HMOD group) had impaired relaxation (E/A <0.8), representing Grade I diastolic dysfunction. This distinction is clinically important for hypertension management: impaired relaxation is common in ageing and hypertension but does not necessarily indicate elevated filling pressures. True diastolic dysfunction with elevated filling pressures — a driver of hypertension-related heart failure — was uncommon in this relatively young cohort.

### Poor Performance of Imported Risk Calculators for Hypertension Treatment Decisions

Hypertension specialists use ASCVD risk scores to guide decisions about intensifying antihypertensive therapy. Agyekum et al.^9^ recently demonstrated that four commonly used ASCVD risk calculators: the Pooled Cohort Equation,^14^ Framingham Risk Score,^15^ WHO/ISH charts,^16^ and Globorisk^17^, show only poor-to-moderate agreement in the Ghanaian population (kappa 0.3–0.8). The WHO/ISH tool, recommended by Ghana’s national CVD guidelines,^10^ classified 82% of adults aged 40–74 as low risk. Our finding that the PCE performed at chance level (AUC 0.466) for detecting multidomain HMOD corroborates this concern. Relying on these scores to guide blood pressure treatment intensity in West Africa would systematically undertreat high-risk individuals with established HMOD.

The clinical implication is serious: using the WHO/ISH tool as currently recommended may undertriage many Ghanaians who are at high risk, leading to missed opportunities for primary prevention.

### baPWV as a Pragmatic Tool for HMOD Detection in Hypertensive Adults

In hypertensive adults, baPWV identifies HMOD burden that conventional risk scores miss entirely. The OMRON/Colin VP-2000 device^21^ is non-invasive, relatively inexpensive compared with echocardiography, and can be operated by trained technicians in community settings. The robustness of this finding was confirmed in a sensitivity analysis using a stricter HMOD threshold (≥3 domains): baPWV remained a strong independent predictor (OR 1.77, 95% CI 1.20–2.62, p=0.004; AUC 0.874; Table S2). The 2023 ESC Guidelines recommend baPWV as a marker of subclinical atherosclerosis (Class IIb).^20^ Our data extend this recommendation to hypertension management in sub-Saharan Africa, where traditional risk factors may not capture the full burden of HMOD.

### Comparison with Other SSA and Global Studies

The prevalence of multidomain HMOD in Ghana (21%) is comparable to or higher than reported in other SSA settings. In the RODAM study of Ghanaian migrants in Europe, the prevalence of any TOD was 17.5% among those living in Ghana versus 25.6% among migrants in Amsterdam.^22^ Direct comparison is limited by differences in TOD definition. In the CARRS study in South Asia, the prevalence of subclinical TOD was 18.5% among adults with diabetes.^23^

Compared with high-income countries, Ghana has a younger age distribution. In the USA, the prevalence of subclinical CVD (coronary artery calcium >0) among adults aged 45–54 years is approximately 50%.^24^ The burden in Ghana is substantial given the younger population and lower levels of risk factor treatment.^25^

### Clinical Implications for Hypertension Care in West Africa

1. **Hypertension control:** Blood pressure control must remain the top priority; 73% of those with multidomain HMOD were hypertensive. Target <130/80 mmHg (or <140/90 mmHg where resources are limited). Ghana’s national guidelines recommend thiazide/CCB-based therapy.^10,25^
2. **HMOD screening using baPWV:** In hypertensive adults aged ≥40 years, baPWV can identify those with HMOD burden missed by conventional risk scores. The number needed to screen is 5 overall and only 2 in adults aged ≥60 years.
3. **Lifestyle interventions:** Population-wide campaigns should promote physical activity and traditional diets rich in vegetables, legumes, and fish.^26^
4. **Health system strengthening:** Training CHPS officers to measure blood pressure, use baPWV where available, and initiate low-cost antihypertensive therapy could substantially reduce HMOD burden.^29^
5. **Research priorities:** Prospective studies are needed to derive and validate a Ghana-specific risk model incorporating baPWV to guide antihypertensive treatment intensification.

### Limitations

Several limitations should be acknowledged. First, the cross-sectional design precludes causal inference. Second, the multidomain HMOD composite, while based on established guidelines,^27^ is a surrogate endpoint that requires prospective validation against hard cardiovascular events. Third, UACR data were missing in 36.8% of participants, which may have underestimated the renal domain contribution. Fourth, the baPWV device (OMRON/Colin VP-2000) was not specifically validated in Ghanaian or West African populations, although it has been extensively used and validated in Asian and European cohorts.^21^ Fifth, our sample was predominantly urban (80.4%), which may limit generalizability to rural settings. Finally, behavioural risk factors were self-reported and subject to recall bias.

### Strengths

Despite these limitations, this study has major strengths. It is among the first to describe multidomain HMOD in a geographically diverse sub-Saharan African population using comprehensive, multi-modal phenotyping (vascular, cardiac, ECG, valvular, renal). The large sample size (N=1,106) and rigorous sampling strategy ensure reasonable generalizability to the Ghanaian adult population. The comparison of multiple individual markers within the same participants provides a unique opportunity to identify the most efficient HMOD assessment tool for resource-limited hypertension care.

### Perspectives

Cardiovascular risk stratification in sub-Saharan Africa remains dependent on Western-derived equations that perform no better than chance for detecting multidomain hypertension-mediated organ damage (HMOD) (AUC 0.466). Brachial-ankle pulse wave velocity — measurable with the same device used for blood pressure, achieves AUC 0.827, 95% CI 0.794–0.860; ΔAUC +0.351, DeLong test p<0.001), recovering prognostic information that conventional risk scoring misses entirely.

The steep age gradient (8.6% TOD prevalence under 45 years rising to 44.4% at ≥60, NNS of 2) defines a practical intervention window. Adults aged 45–59 years are the highest-yield screening group: old enough to carry substantial subclinical burden, yet young enough that blood pressure control and lifestyle modification can alter trajectory.

Two priorities emerge: a Ghana-specific ASCVD risk model incorporating baPWV and ABI must be validated against hard endpoints in a prospective cohort; and geographic clustering of carotid plaque in the Ashanti region warrants investigation for modifiable exposures with implications for screening programmes across West Africa.

## Conclusion

One in five Ghanaian adults has hypertension-mediated organ damage in two or more organ systems, with prevalence increasing sharply from 8.6% under age 45 to 44.4% at age ≥60. baPWV, measurable in minutes with a single automated device — is the single strongest predictor and demonstrated superior discriminative performance (AUC 0.827, 95% CI 0.794–0.860) compared with the ASCVD Pooled Cohort Equations (AUC 0.466; ΔAUC +0.351, DeLong test p<0.001). Hypertension is the central driver, present in 73% of TOD-positive adults. Although our multidomain HMOD composite is a surrogate endpoint requiring future validation against hard clinical outcomes, it provides a clinically meaningful snapshot of systemic subclinical disease burden. The striking discrepancy between ECG-LVH (42%) and echocardiographic LVH (6.4%) cautions against reliance on ECG criteria alone in Black African populations. There is an urgent need for targeted prevention focusing on hypertension control and lifestyle modification, alongside development of locally derived risk models incorporating baPWV.

### Novelty and Relevance

#### What Is New?

- One of the first comprehensive multi-domain TOD assessments in a West African community cohort.
- baPWV demonstrated superior discriminative performance (AUC 0.827 vs. 0.466; ΔAUC +0.351, DeLong test p<0.001).

#### What Is Relevant?

- 21.2% of Ghanaian adults have subclinical damage in ≥2 organ systems; prevalence reaches 44.4% at age ≥60.
- Hypertension drives 73% of TOD cases; baPWV is the strongest single predictor (NNS = 5 overall, 2 for ≥60 years).

#### Clinical Implications

- baPWV should supplement risk scoring in hypertensive adults ≥45 years across West Africa.
- ECG voltage criteria have poor specificity for LVH in Black Africans (42% ECG-LVH vs. 6.4% echo-LVH).

## Data Availability

The de-identified dataset used in this secondary analysis is available from the corresponding author upon reasonable request and subject to institutional data transfer agreements.

## Author contributions (CRediT)

K.O. Agyapong: Conceptualization, Methodology, Formal Analysis, Writing – Original Draft, Writing – Review & Editing.

E. Kyeremah: Data Curation, Writing – Review & Editing.

A.A. Folson: Resources, Data Curation, Writing – Review & Editing.

F. Agyekum: Resources, Data Curation, Writing – Review & Editing.

K. R. M. Blenman: conceptualisation, Writing – Review & Editing.

L.T. Appiah: Data Curation, Writing – Review & Editing.

Y. Adu-Boakye: Data Curation, Writing – Review & Editing.

I.K. Owusu: Conceptualization, Resources, Supervision, Writing – Review & Editing.

## Disclosure

## Data availability

The datasets analysed in this study are part of the Ghana Heart Study. De-identified participant-level data may be requested from the corresponding author, subject to institutional data governance requirements and a standard data transfer agreement approved by Ghanaian institutions and Guangdong Provincial People’s Hospital.

## Clinical Trial Registration

Chinese Clinical Trial Registry: **ChiCTR1800017374**. URL: http://www.chictr.org.cn/showprojen.aspx?proj=30439

## Acknowledgments

The authors would like to express their sincere gratitude to all the participants who were involved in this study, and the research assistants for the support they provided. Without their cooperation, this study would not have been successful.

Supplementary information is available at http://www.oup.com/ajh

## Supplementary Tables

**Table S1.**
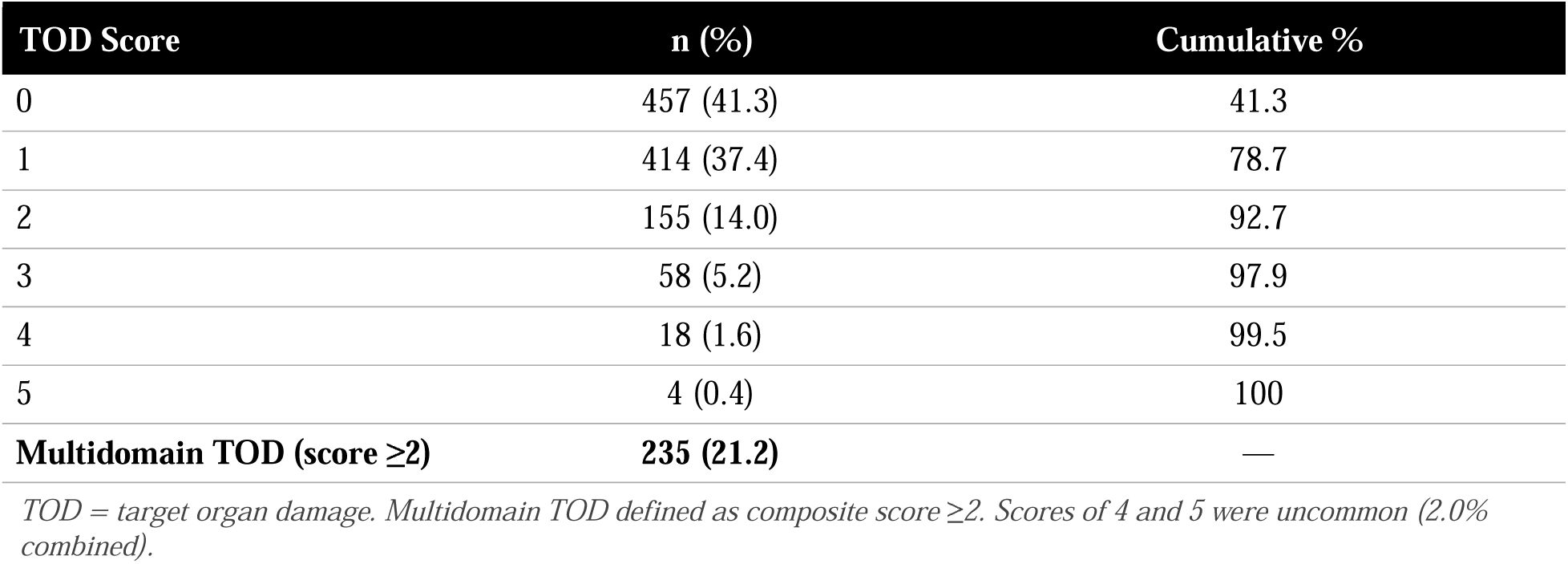
Distribution of the TOD Composite Score (N=1,106)

| TOD Score | n (%) | Cumulative % |
| --- | --- | --- |
| 0 | 457 (41.3) | 41.3 |
| 1 | 414 (37.4) | 78.7 |
| 2 | 155 (14.0) | 92.7 |
| 3 | 58 (5.2) | 97.9 |
| 4 | 18 (1.6) | 99.5 |
| 5 | 4 (0.4) | 100 |
| **Multidomain TOD (score $\geq 2$)** | **235 (21.2)** | — |

TOD = target organ damage. Multidomain TOD defined as composite score $\geq 2$ . Scores of 4 and 5 were uncommon (2.0% combined).

**Table S2.** Sensitivity Analysis: Stricter TOD Threshold (≥3 Domains)

| Parameter | TOD $\geq 2$ (Primary) | TOD $\geq 3$ (Sensitivity) |
| --- | --- | --- |
| Prevalence, n (%) | 235 (21.2) | 80 (7.2) |
| baPWV OR (per SD, 95% CI) | 1.44 (1.29–1.60) | 1.77 (1.20–2.62) |
| p-value | <0.001 | 0.004 |
| baPWV AUC (95% CI) | 0.827 (0.794–0.860) | 0.874 (0.839–0.909) |

OR = odds ratio; SD = standard deviation; AUC = area under the curve; baPWV = brachial-ankle pulse wave velocity. The primary threshold ( $\geq 2$ domains) and the sensitivity threshold ( $\geq 3$ domains) are compared. baPWV remained a significant predictor at both thresholds. Adjusted for age, systolic BP, BMI, dyslipidaemia, and smoking.

**Table S3.** Incremental Discriminative Value of baPWV Over the ASCVD Pooled Cohort Equations.

| Model | AUC | ΔAUC vs. ASCVD alone | p-value (DeLong) |
| --- | --- | --- | --- |
| ASCVD alone | 0.466 | Reference | — |
| ASCVD + baPWV | 0.827 | +0.351 | <0.001 |

AUC = area under the receiver operating characteristic curve; ΔAUC = change in AUC; ASCVD = atherosclerotic cardiovascular disease; baPWV = brachial-ankle pulse wave velocity. ΔAUC was calculated using DeLong's method for paired ROC curves. Likelihood ratio test: deviance = 764.28, $p < 2.2 \times 10^{-16}$ .

**Table S4.** Three-Domain Cardiometabolic Triad Analysis.

| Triad | Definition | n (%) | Age-Std % (SE) | Adjusted OR (95% CI) | p |
| --- | --- | --- | --- | --- | --- |
| Metabolic Syndrome | Central obesity + hypertension + (dyslipidaemia OR DM) | 148 (13.4) | 12.6 (0.94) | 2.11 (1.28–3.49) | 0.004 |
| Vascular-Renal | High baPWV + low ABI + CKD stage $\geq 3$ | 2 (0.2) | 0.15 (0.10) | Not estimable | 0.98 |
| Cardio-Renal | Echo-LVH + CKD stage $\geq 3$ + hs-CRP $\geq 3$ mg/L | 4 (0.4) | 0.31 (0.16) | Not estimable | 0.97 |

OR = odds ratio; CI = confidence interval; DM = diabetes mellitus; ABI = ankle-brachial index; CKD = chronic kidney disease; Echo-LVH = echocardiographic left ventricular hypertrophy; hs-CRP = high-sensitivity C-reactive protein. Age-standardised prevalence calculated using the direct method with the study population as the reference. Adjusted for age, systolic BP, BMI, dyslipidaemia, and smoking. Vascular-renal and cardio-renal triads were too rare for logistic regression.

